# Development and Application of Pharmacological Statin-Associated Muscle Symptoms Phenotyping Algorithms Using Structured and Unstructured Electronic Health Records Data

**DOI:** 10.1101/2023.05.04.23289523

**Authors:** Boguang Sun, Pui Ying Yew, Chih-Lin Chi, Meijia Song, Matt Loth, Rui Zhang, Robert J. Straka

## Abstract

**Background:** Statins are widely prescribed cholesterol-lowering medications in the US, but their clinical benefits can be diminished by statin-associated muscle symptoms (SAMS), leading to discontinuation. In this study, we aimed to develop and validate a pharmacological SAMS clinical phenotyping algorithm using electronic health records (EHRs) data from Minnesota Fairview.

**Methods:** We retrieved structured and unstructured EHR data of statin users and manually ascertained a gold standard set of SAMS cases and controls using the SAMS-CI tool from clinical notes in 200 patients. We developed machine learning algorithms and rule-based algorithms that incorporated various criteria, including ICD codes, statin allergy, creatine kinase elevation, and keyword mentions in clinical notes. We applied the best performing algorithm to the statin cohort to identify SAMS.

**Results:** We identified 16,889 patients who started statins in the Fairview EHR system from 2010-2020. The combined rule-based (CRB) algorithm, which utilized both clinical notes and structured data criteria, achieved similar performance compared to machine learning algorithms with a precision of 0.85, recall of 0.71, and F1 score of 0.77 against the gold standard set. Applying the CRB algorithm to the statin cohort, we identified the pharmacological SAMS prevalence to be 1.9% and selective risk factors which included female gender, coronary artery disease, hypothyroidism, use of immunosuppressants or fibrates.

**Conclusion:** Our study developed and validated a simple pharmacological SAMS phenotyping algorithm that can be used to create SAMS case/control cohort for further analysis such as developing SAMS risk prediction model.

**LAY SUMMARY:** Statins are commonly prescribed cholesterol-lowering medications in the US, but some patients may experience statin-associated muscle symptoms (SAMS) that can reduce their benefits. In this study, we developed and tested a simple algorithm using electronic health records (EHRs) to identify cases of SAMS. We retrieved data from statin users in the Minnesota Fairview EHR system and manually identified a gold standard set of SAMS cases and controls using a clinical tool. We developed machine learning and rule-based algorithms that considered various criteria, such as ICD codes, statin allergy, creatine kinase elevation, and keyword mentions in clinical notes. The best performing algorithm, called the combined rule-based (CRB) algorithm, achieved similar performance to machine learning algorithms in identifying SAMS cases. When applied to the larger statin cohort, the CRB algorithm identified a prevalence of 1.9% for pharmacological SAMS, and identified selective risk factors such as female gender, coronary artery disease, hypothyroidism, and use of immunosuppressants or fibrates. The developed algorithm has the potential to help create SAMS case/control cohorts for future studies such as building models to predict SAMS risks for patients.

## INTRODUCTION

Nearly half of Americans over 65 years of age take statins, a class of cholesterol lowering medications proven to reduce morbidity and mortality.^1^ However, around 25 to 50% of statin users do not fully experience the benefits of statins because of statin discontinuation.^2^ Among the reasons for statin discontinuation are personal preference, financial burdens, or side effects. Around 25% former statin users attributed their non-adherence or discontinuation to side effects, predominantly statin-associated muscle symptoms (SAMS).^2^ Post-market pharmacovigilance of adverse drug reactions (ADRs) including SAMS, are crucial to ensure that medications are safe in the long term. FDA Adverse Event Reporting System is a well-recognized safety surveillance program for all approved medications and therapeutic biologics.^3^ However, studies have found underreporting of certain ADRs in the FAERS dataset compared to the real-world evidence.^4,5^ Furthermore, clinicians might not routinely report certain ADRs to the FDA, especially when they are familiar or insidious, as is often the case with SAMS. Therefore, in order to optimize the appropriate use of these life-saving medications, there is a critical need to identify the predictors of the development of SAMS, based on real-world data where there is sufficient documentation of longitudinal use.

In recent years, with the increasing usage of Electronic Health Records (EHRs) as patient data warehouses, targeted mining of real-world data stored in EHRs has garnered attention as an alternative means for ADR detection and monitoring.^6^ In EHRs, 20% of patient-centered data is in structured format such as procedures and laboratory tests whereas 80% takes the form of unstructured data consisting of clinical notes in the free-text format.^6^ Signals within EHRs that offer evidence for SAMS manifestations include International Classification of Diseases (ICD) coding of muscle symptoms such as myopathy and myalgia, patients’ allergy list specific to statin intolerance, temporal creatine kinase (CK) elevation and most importantly, clinicians’ notes documenting the incidence and development of SAMS during patient visits.

To date, SAMS clinical phenotyping algorithms developed based on various EHR systems have shown that a combination of structured and unstructured SAMS-related EHR signals can better identify SAMS compared to using structured data alone.^7,8^ However, the cross-institution generalizability of such algorithms is unknown. Furthermore, current studies have not investigated the specific phenotyping of pharmacological SAMS (non-nocebo SAMS). To that end, we aim to develop and validate a pharmacological SAMS clinical phenotyping algorithm based on the University of Minnesota’s (UMN) Clinical Data Repository (CDR) with a coverage of Fairview EHRs which includes information from six hospitals and over 115 clinics within Minnesota. We applied a scalable NLP-PIER (Natural Language Processing-Patient Information Extraction for Research) tool integrated within the EHR database to search for clinical notes associated with SAMS.^9^ We utilized the validated SAMS-Clinical Index (SAMS-CI) tool to ascertain pharmacological SAMS and develop gold standard manual annotations for our phenotyping algorithm.^10^ We also applied the best-performing phenotyping algorithm to classify the SAMS (case) and non-SAMS (control) cohorts and reported the differences in patient characteristics and risk factors associated with SAMS. These identified case and control cohorts can be utilized to develop pharmacological SAMS risk prediction models based on clinical features extracted from the EHRs.

## METHODS

### Data source and cohort identification

We retrieved our study cohort from Fairview EHR between 1/1/2010 to 12/31/2020, which represents our study period. As shown in Figure 1, the overall statin cohort contains patients over 18 years old at index date and were regular Fairview system users. We defined regular Fairview system users as having at least one record of each of the following during both the baseline and follow-up periods: 1) Fairview encounter, 2) blood pressure or weight measurements, 3) Fairview pharmacy dispensing, and 4) laboratory data.^11^

**Figure 1:**
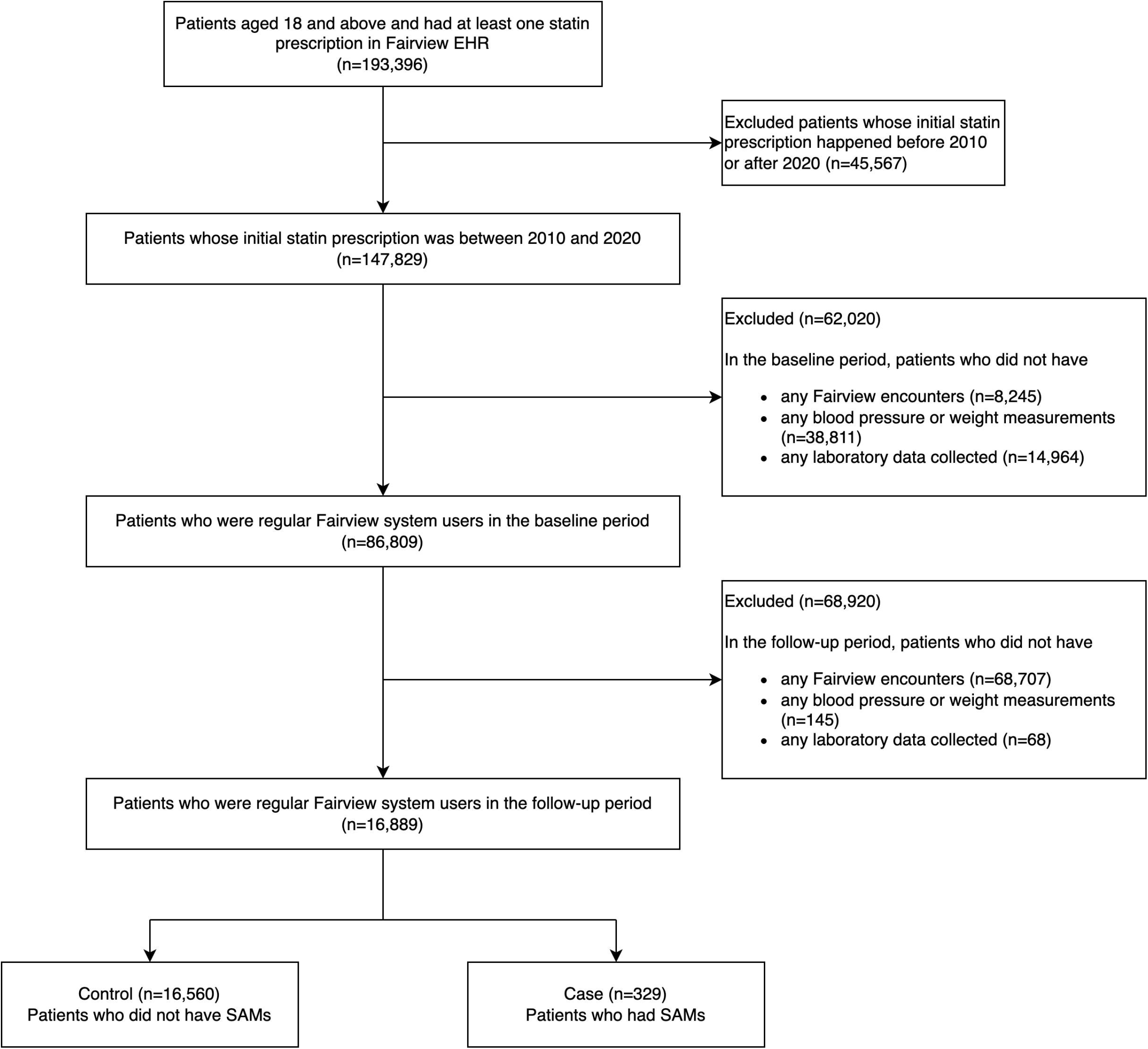
Study Flowchart for Statin Cohort Selection from Fairview EHR

Index date was the day the patient was prescribed their first statin medications (atorvastatin, fluvastatin, lovastatin, pitavastatin, pravastatin, rosuvastatin and simvastatin). The baseline period used to define demographic, comorbidity and social history was a year preceding the index date. The baseline period to define co-medications was three months preceding the index date. The follow-up period was 1 year after the index date or the end of the study period, whichever was earlier.

For the statin cohort, we included patients who initiated any statins and were regular Fairview EHR users during the study period. To exclude prevalent statin users, we excluded any patient who had any statin prescriptions prior to the index date.

We retrieved and analyzed structured EHR data within the relational databases containing patient demographics, medications, laboratory and procedures maintained by UMN Academic Health Center-Information Exchange (AHC-IE) team. We obtained and searched for clinical notes related to SAMS using the NLP-PIER tool, an NLP search engine enabled by AHC-IE.^9^ Figure 2 demonstrates the overview of study workflow and methodology. The study is approved by University of Minnesota IRB (STUDY00011134).

**Figure 2:**
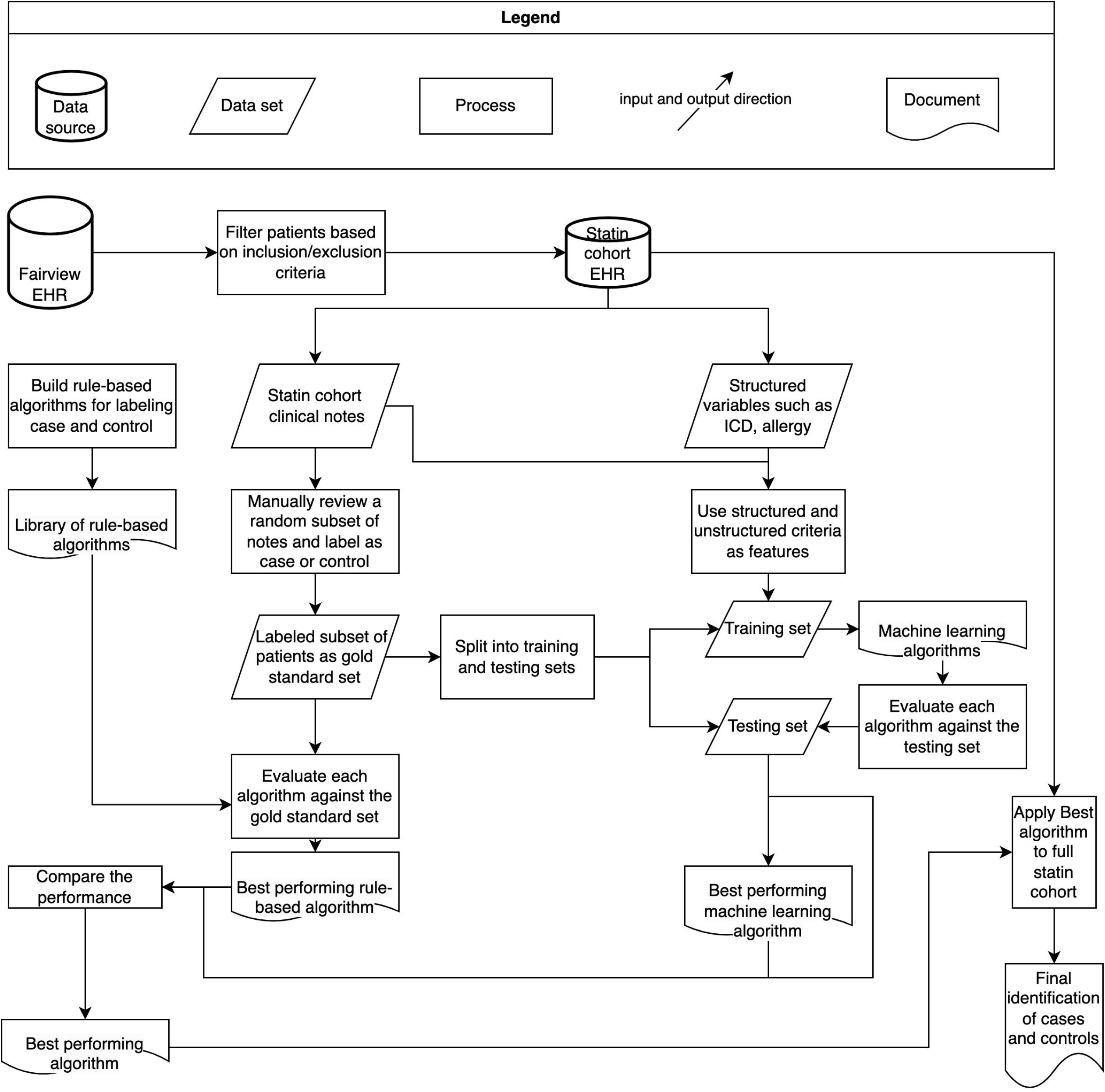
Overview of Workflow and Methodology

### Manual case ascertainment

To examine the structure and documentation styles of clinical notes within Fairview EHR, we randomly selected clinical notes from 100 patients where their notes included any mentioning of a named statin medication 10 words before or after mentioning reference to any muscle complaints such as muscle pain, myalgia or myopathy after the index date. Then, we created the NLP-PIER search term that includes mentions of statin medications, muscle symptoms and excludes the negation phrases such as “no myalgia” or “deny myopathy”. Next, we created the gold standard set using clinical notes from another independent 200 patients. These 200 patients consisted of a balanced number of potential SAMS cases and non-SAMS controls classified by the NLP-PIER search term.

Two domain experts (with either pharmacy or nursing backgrounds) manually reviewed and annotated the clinical notes in the gold standard set. The manual reviewers annotated and ascertained the pharmacological SAMS cases based on the SAMS-CI tool.^10^ This tool aims to discern pharmacological SAMS from nocebo SAMS by incorporating muscle distribution of symptoms, temporal patterns (symptom onset after statin initiation, improvement and recurrence after statin discontinuation and rechallenge) into a scoring system. This tool was also prospectively utilized to ascertain SAMS in an ongoing clinical trial.^12^

In Table S1, we showed our use case of the SAMS-CI scoring symptoms and clinical scenarios for each score assignment. In Table S2, we provided two case vignettes in supplement to further illustrate how we determined the scores for each patient.

Overall, we assigned patients with a score greater or equal to 7 points as SAMS cases according to the SAMS-CI tool.^10^ We used Cohen’s kappa values to assess the manual review agreement between the reviewers in the gold standard set.

### Algorithm development: rule-based and machine learning (ML) algorithms

We considered six rule-based algorithms: 1) ICD codes only; 2) allergy list only; 3) CK elevation only; 4) ICD codes or allergy list or CK elevation; 5) clinical notes mentions only and 6) combination of structured data (ICD codes, allergy list and CK elevation) and clinical notes mentions. The follow-up period for each individual criterion (ICD codes, allergy list, CK elevation, and clinical notes mentions) was one year after the index date. We selected a one-year follow-up because this was the timeframe that most statin adverse events occur.^13^

ICD criterion: Table S3 shows the specific ICD codes we included as signals for SAMS. Patients met the ICD criterion if they only had ICD codes after the index date (no prior ICD codes documentation of muscle symptoms).

Allergy criterion: Patients met allergy criterion if their allergy list in the EHR indicated having muscle symptoms due to statin medications.

CK elevation criterion: We chose to use a threshold of CK > 3 times the upper limit of normal according to the SEARCH (Study of the Effectiveness of Additional Reductions in Cholesterol and Homocysteine) Trial.^14^ The CK normal ranges used were 30 to 145 U/L for females and 55 to 170 U/L males.^15^

Clinical notes mentions criterion: patients met the notes mentioning criterion if, after the index date, there were any mentions of statin medications 10 words before and after the mentioning of muscle complaints without mentions of negation phrases (NLP-PIER search term).

Combined rule-based (CRB) algorithm: Figure 3 shows the decision flowchart for the pharmacological SAMS identification CRB algorithm.

**Figure 3:**
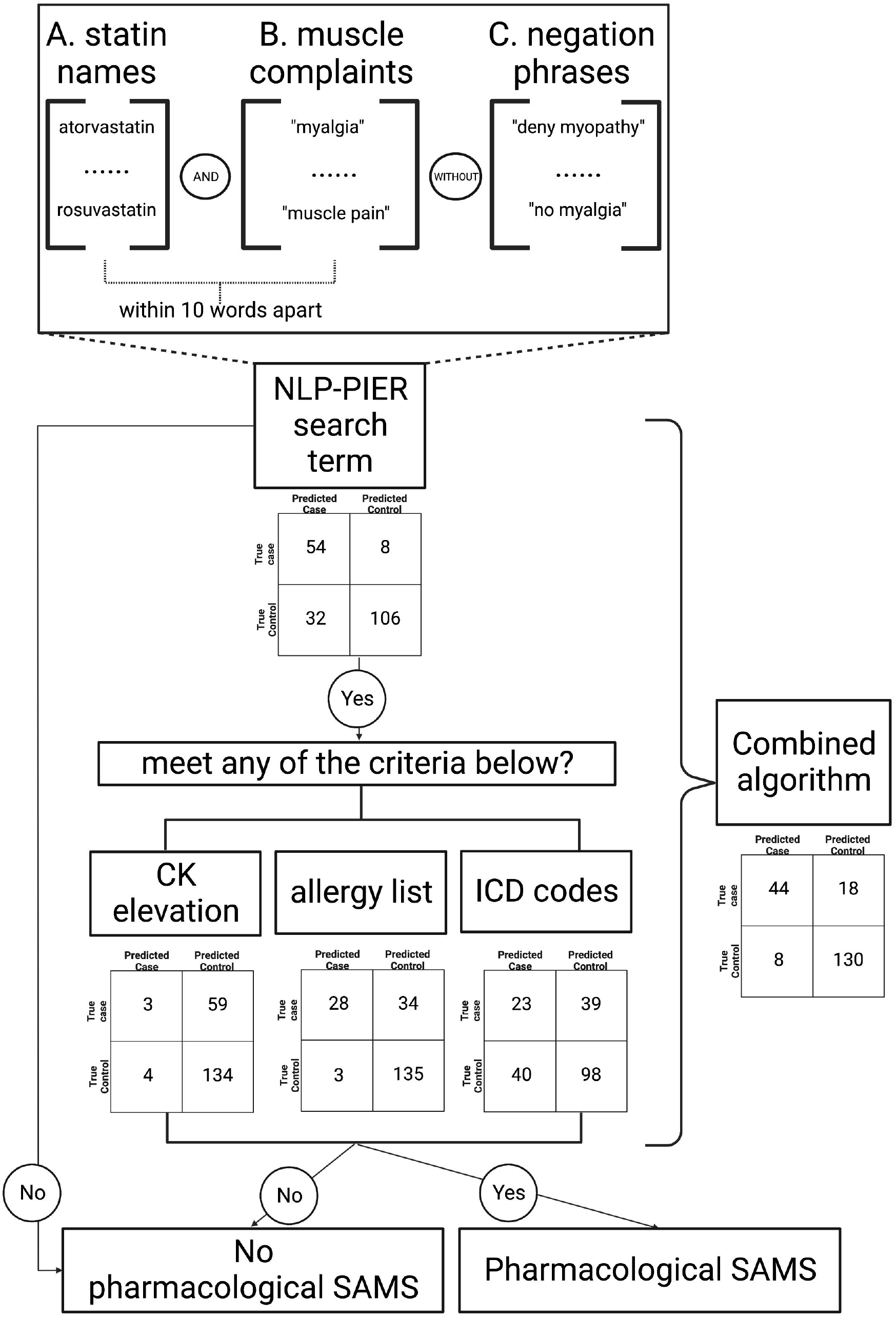
Flowchart for the Individual and Combined Rule-Based (CRB) Algorithms

For the ML algorithms, we used common ML classifiers including Decision Tree (DT), Support Vector Machine (SVM), K-Nearest Neighbors (KNN), Random Forest (RF), AdaBoost (AB). We split the gold standard set into 70% training and 30% testing set. We used the four rule-based labels (ICD codes, allergy list, CK elevation and clinical notes mentions) as binary features to train the ML classifiers.

### Algorithm evaluation and application

We evaluated the rule-based algorithms against the whole gold standard set. Next, we compared the best-performing rule-based algorithm with ML algorithms against the whole gold standard set (also referred to as the overall set) and gold standard testing set (referred to as the testing set thereafter). The gold standard testing set is a subset within the whole gold standard set as specified in the previous section. We reported the performance of each algorithm (precision, recall and F1 scores) in this binary classification problem to separate patients into cases (have pharmacological SAMS) or controls (do not have pharmacological SAMS).

Then, we applied the best-performing algorithm to the overall statin cohort. We reported preliminary patient baseline characteristics between cases and controls including demographics, social history, comorbidities, and concurrent medications associated with pharmacological SAMS risk.^16^ We conducted univariate and multivariate association analysis of pharmacological SAMS outcome and risk factors. We included statistically significant (p<0.05) baseline factors from the univariate analysis in the multivariate analysis.

## RESULTS

### Cohort identification

As shown in Figure 1, out of the 193,396 adult patients who started statins in the Fairview EHR system, we included 16,889 patients who met our criteria in this study: patients who started statins during 2010 to 2020, were regular Fairview EHR users, and were not prevalent statin users.

### Manual case ascertainment

Two reviewers annotated clinical notes from 200 patients in the gold standard set. The two reviewers achieved high agreement in determining the case vs controls using the SAMS-CI tool (kappa = 0.985).

In the gold standard set, the NLP-PIER search term identified 86 cases and 114 controls. After manual reviews, we ascertained 62 cases and 138 controls (true cases and true controls).

### Algorithm development and evaluation

Figure 3 shows the flowchart for the CRB algorithm. Specifically, the CRB algorithm determined the patient to have pharmacological SAMS when they met (1) the clinical notes mentions criterion and; (2) at least one of the structured data criteria (ICD codes, CK elevation or allergy list). As demonstrated in Figure 3, the NLP-PIER search term for SAMS has three components: A. mentioning of any statin medications (see methods for the statin medications list); B. mentioning of any muscle complains including “myalgia”, “myopathy”, “muscle pain”, “muscle ache”, “muscle cramp”, “myositis”; C. with the mentioning of negation phrases including “no myalgia”, “no myopathy”, “deny myalgia”, “deny myopathy”, “suspect myalgia”, “no muscle aches”, “monitor for myalgia”. The mentioning of criteria A and B has to be within 10 words apart. In Figure 2, we also reported the algorithm performance in individual and CRB algorithms using the confusion matrices from the gold standard set (200 patients).

As shown in Table 1, the CRB algorithm achieved better performances compared to the other rule-based algorithms. The precision, recall and F1 score were 0.85, 0.71, 0.77 against the gold standard set (N=200), respectively. The ICD only algorithm had the worst performance compared with other algorithms with an F1 score of only 0.37 against the gold standard set. The allergy only algorithm had good performance in terms of precision (0.90 against the gold standard set) but its recall was compromised (0.45 against the gold standard set). The notes-only algorithm had better performance in terms of recall compared to the CRB algorithm but was outperformed by the CRB algorithm regarding the precision (0.63 against the gold standard set).

**Table 1:** Rule-based Algorithm Performances by Precision, Recall and F1 Scores

|  | Precision | Recall | F1 score |
| --- | --- | --- | --- |
| ICD only | 0.37 | 0.37 | 0.37 |
| Allergy only | 0.90 | 0.45 | 0.60 |
| CK elevation only | 0.43 | 0.05 | 0.09 |
| ICD or allergy or CK elevation | 0.53 | 0.77 | 0.63 |
| Notes only | 0.63 | 0.87 | 0.73 |
| **Combined rule-based algorithm** | **0.85** | **0.71** | **0.77** |

As shown in Table 2, the CRB rule-based algorithm achieved similar performances compared to the ML algorithms in the overall set (N=200) and testing set (N=60). The ML classifiers such as RF and AB had slightly better recall than the CRB algorithm when evaluating against the overall set but the differences in recall were diminished when compared against the testing set.

**Table 2.** Combined rule-based and Machine Learning Algorithm Performances

| Algorithms | Precision | Recall | F1 score |
| --- | --- | --- | --- |
| **Combined rule-based** |  |  |  |
| **Overall set (N=200)** | **0.85** | **0.71** | **0.77** |
| **Testing set (N=60)** | **0.89** | **0.84** | **0.86** |
| Random forest |  |  |  |
| Overall set | 0.80 | 0.76 | 0.78 |
| Testing set | 0.80 | 0.84 | 0.82 |
| Adaptive boosting |  |  |  |
| Overall set | 0.82 | 0.74 | 0.78 |
| Testing set | 0.84 | 0.84 | 0.84 |
| K-nearest neighbors |  |  |  |
| Overall set | 0.84 | 0.68 | 0.75 |
| Testing set | 0.88 | 0.79 | 0.83 |
| Decision tree |  |  |  |
| Overall set | 0.90 | 0.45 | 0.60 |
| Testing set | 0.82 | 0.47 | 0.60 |

### Algorithm applications: patient characteristics comparison and association analysis

After applying the CRB algorithm to the statin cohort, we identified 329 cases and 16,560 controls. This translated to a pharmacological SAMS prevalence of 1.9% (329/16,889) in our statin cohort. Table 3 shows the baseline characteristics of the cases and controls. Briefly, the mean age was 67.1 vs 66.8 in cases and controls, respectively. The pharmacological SAMS case group had significantly more females than the controls (50.5 vs 44.5%, p<0.05). Additionally, the SAMS cases group had significantly more hypertension (74.2 vs 66.3%), coronary artery disease (52.9 vs 37.3%), chronic kidney disease (11.9 vs 7.6%), and hypothyroidism (20.1 vs 12.8%) than the controls. Significantly more patients in the cases group took beta-blockers (53.5 vs 45%), immunosuppressants (13.7 vs 8.3%), and fibrates (4.5 vs 2.2%) than the controls.

**Table 3:** Demographic and Baseline Characteristics of the pharmacological SAMS Case and Control Patients^1^

|  | Case (N=329) | Control (N=16,560) | p values |
| --- | --- | --- | --- |
| Age — year | 67.1 ± 12.8 | 66.8 ± 13.9 | 0.58 |
| Female sex | 166 (50.5) | 7374 (44.5) | **0.04<sup>2</sup>** |
| Race<sup>3</sup> |  |  |  |
| White | 296 (89.9) | 14489 (87.5) | 0.6 |
| Asian | 7 (2.1) | 385 (2.3) |  |
| Black | 15 (4.6) | 854 (5.2) |  |
| Other | 11 (3.4) | 832 (5) |  |
| Body-mass index — kg/m<sup>2</sup> | 30.1 ± 6.5 | 29.9 ± 7.6 | 0.55 |
| High intensity statins |  |  |  |
| Social history |  |  |  |
| Smoking | 98 (29.8) | 5403 (32.6) | 0.3 |
| Alcohol | 150 (45.6) | 7121 (43) | 0.38 |
| Medical history |  |  |  |
| Hypertension | 244 (74.2) | 10985 (66.3) | **<0.01** |
| Diabetes mellitus | 105 (31.9) | 5121 (30.9) | 0.75 |
| Coronary artery disease | 174 (52.9) | 6176 (37.3) | **<0.01** |
| Congestive heart failure | 78 (23.4) | 3360 (20.3) | 0.15 |
| Chronic Kidney Disease | 39 (11.9) | 1257 (7.6) | **<0.01** |
| Hypothyroidism | 68 (20.1) | 2122 (12.8) | **<0.01** |
| Medication history |  |  |  |
| Angiotensin-converting enzyme inhibitors | 101 (30.1) | 4579 (27.7) | 0.25 |
| Beta-blockers | 176 (53.5) | 7458 (45) | **<0.01** |
| Immunosuppressants<sup>4</sup> | 45 (13.7) | 1367 (8.3) | **<0.01** |
| Fibrates | 14 (4.5) | 370 (2.2) | **0.02** |

<sup>1</sup>Baseline characteristics were defined within one year preceding the index date.
<sup>2</sup>Bolding denotes statistical significance.
<sup>3</sup>Categorical variables in count (%) while continuous variables in mean ( ± standard deviation).
<sup>4</sup>Immunosuppressants include cyclosporine, everolimus, sirolimus, tacrolimus.
<sup>5</sup>Fibrates include fenofibrates and gemfibrozil.

As shown in Table 4, all the baseline factors shown to be significantly different between cases and controls were also significant risk factors identified using univariate analysis. However, only female gender, coronary artery disease, hypothyroidism, use of immunosuppressant and fibrates were associated with higher risk of SAMS after the multivariate analysis.

**Table 4.** Univariate and Multivariate Logistic Regression of Risk Factors and Pharmacological SAMS Outcome

|  | Univariate |  | Multivariate |  |
| --- | --- | --- | --- | --- |
|  | OR (95% CI)<sup>1</sup> | p values | OR (95% CI) | p values |
| Age | 1 (0.99, 1.01) | 0.6 |  |  |
| Female sex | **1.27 (1.02, 1.59)** | **0.033** | **1.33 (1.05, 1.67)** | **0.02** |
| Body-mass index | 0.89 (0.82, 1.21) | 0.46 |  |  |
| Smoking | 0.88 (0.69, 1.11) | 0.3 |  |  |
| Alcohol use | 1.11 (0.89, 1.38) | 0.3 |  |  |
| Hypertension | **1.46 (1.14, 1.88)** | **0.003** | 1.26 (0.98, 1.63) | 0.08 |
| Diabetes mellitus | 1.05 (0.83, 1.32) | 0.7 |  |  |
| Coronary artery disease | **1.89 (1.52, 2.35)** | **<0.001** | **1.84 (1.47, 2.32)** | **<0.001** |
| Congestive heart failure | 1.22 (0.94, 1.57) | 0.13 |  |  |
| Chronic Kidney Disease | **1.64 (1.15, 2.27)** | **0.004** | 1.28 (0.88, 1.79) | 0.18 |
| Hypothyroidism | **1.77 (1.34, 2.31)** | **<0.001** | **1.59 (1.19, 2.09)** | **<0.001** |
| Angiotensin-converting enzyme inhibitors | 1.1 (0.98, 1.23) | 0.62 |  |  |
| Beta-blockers | **1.4 (1.13, 1.75)** | **0.002** | 1.16 (0.92, 1.46) | 0.2 |
| Immunosuppressants | **1.76 (1.26, 2.4)** | **<0.001** | **1.66 (1.18, 2.28)** | **<0.001** |
| Fibrates | **1.94 (1.08, 3.23)** | **0.017** | **1.93 (1.07, 3.22)** | **0.02** |

<sup>1</sup>OR is Odds Ratio, CI is Confidence Interval

## DISCUSSION

Studies^17,18^ have demonstrated that EHRs can be used as a reliable source for ADR phenotyping and downstream research such as pharmacovigilance and genetics studies.^19,20^ SAMS, as an example of ADR, has been challenging for phenotyping due to heterogeneity of symptoms and nocebo effects.^21^ As a result, the prevalence of all SAMS ranges from 5% to 25% but the pharmacological (non-nocebo) SAMS to be only about 1-2%.^16^ To date, multiple studies have proposed SAMS phenotyping algorithms either using structured data alone^22^ or in combination with unstructured clinical notes.^7,8^ However, it is unclear whether such tools have reasonable performances across different institutions. On the other hand, the case ascertainment methods varied in these studies without the use of SAMS-CI tool aimed to identify pharmacological SAMS. Therefore, the motivation of our study was to develop a pharmacological SAMS phenotyping algorithm using EHRs data specifically available in the Fairview Healthcare system.

In this study, we first identified the statin user cohort and defined statin index date, baseline and follow-up periods. These timelines were crucial for us to analyze the temporal relationship between statin use and muscle symptoms and calculate the SAMS-CI score. We also defined regular Fairview EHR users to ensure that the patients included had sufficient longitudinal clinical notes of their system encounters. Specifically, each patient in our cohort had approximately 40 statin-related clinical notes. This allowed us to leverage more information within the clinical notes to sufficiently adjudicate the SAMS cases vs controls using the SAMS-CI tool.

We developed pharmacological SAMS phenotype algorithms using structured and unstructured EHR data in an integrated healthcare system. As demonstrated in Table 1, using structured data components alone or in combination such as ICD coding, allergy list or CK elevation as phenotyping algorithms could not identify pharmacological SAMS with reasonable performance. Using clinical notes mentions as a single criterion for SAMS can achieve similar recalls compared to the CRB algorithm but it did not perform well in terms of precision (high false positive rates). Overall, the CRB algorithm with consideration of patients’ allergy list, ICD coding of muscle symptoms, CK elevation and clinical notes mentions achieved the best performance for pharmacological SAMS identification. We designed the CRB algorithm in a hierarchical structure where we gave the clinical notes mentions criterion more weight in determining the cases but also leveraged the other criteria to help increase the performance. Of note, our hierarchical CRB algorithm had overall similar performances when compared with ML algorithms (Table 2). The ML algorithms such as RF and AB had incremental improvement in recall compared to the CRB algorithm. However, since our end-goal was to use the best-performing phenotyping algorithm to classify SAMS cases and controls, high precision becomes a more desirable metric in our model evaluation. Additionally, the rule-based algorithm also has clinical advantage as it is easier to interpret. Overall, we chose the CRB algorithm as the best-performing algorithm for application.

Our study applied the CRB algorithm on the pre-defined statin cohort (N=16,889) as shown in Figure 2. We estimated the prevalence of pharmacological SAMS to be 1.9% (329/16,889), which was similar to the estimation reported in the current National Lipid Association guidelines.^16^ As shown in Table 3, the prevalence or values of several baseline factors were statistically different between SAMS case (N=329) and control (N=16,560) cohorts. After a univariate/multivariate analysis shown in Table 4, we recognized several key risk factors such as female gender, coronary artery disease, hypothyroidism, use of fibrates and immunosuppressants that were associated with increased risk of pharmacological SAMS in our statin cohort. These risk factors identified in our analysis align with common SAMS risk factors in real-world settings^16^ and also previously recognized in national guidelines^23^ thus further strengthening the potential clinical usability of our phenotyping algorithm.

Our study had some limitations. First of all, the generalizability of our proposed phenotyping algorithm to other EHR systems is unknown. However, we believe our phenotyping algorithm development framework (Figure 2 and 3) might have potential for interoperability among different EHR systems. This is because each individual component in the CRB algorithm is readily available in other EHR systems. We also did not over-train the NLP-PIER search term by adding additional filter words. We intended to make the NLP-PIER search term a “weak learner” and when combined with other features such as ICD codes, CK elevation and allergy list, the model performance was optimized. Secondly, in this study, we focused primarily on rule-based and ML algorithms that utilize EHR components (ICD codes, CK elevation, allergy list and clinical notes mentions) for prediction. We appreciate that novel ML and deep learning NLP approaches leveraging clinical notes might achieve better performances compared to conventional phenotyping algorithms.^24^ Therefore, future studies are needed to develop pharmacological SAMS phenotyping algorithms using novel NLP techniques.

For future steps, we will develop and validate a pharmacological SAMS risk prediction model using the pharmacological SAMS cases and control cohorts classified by our pharmacological SAMS phenotyping algorithm. We envision that the risk prediction model can be incorporated into patients’ EHRs as an element of clinical decision support. Once a patient has an indication for a statin and at the same time, been deemed as high risk for developing SAMS, a “warning or cautionary message” could fire in the EHR prompting a review by prescribers so that preemptive measures (adjustment of doses and selection of specific statin, reviews of interacting medications and more frequent monitoring, etc.) can be taken to improve statin adherence.

## CONCLUSION

In this study, we developed a pharmacological SAMS phenotyping algorithm using structured and unstructured data within the Fairview EHRs. The CRB algorithm incorporating unstructured and structured data outperformed all other rule-based algorithms with precision, recall and F1 of 0.85, 0.71, 0.77 against the gold standard set, respectively. The CRB algorithm also had comparable performances to ML algorithms. We applied the best-performing CRB algorithm on the statin cohort and identified the pharmacological SAMS prevalence of 1.9% and pharmacological SAMS risk factors including female gender, coronary artery disease, hypothyroidism, use of immunosuppressants and fibrates. These observations align with the real-world clinical practice estimates of pharmacological SAMS which further corroborate the clinical utility of our algorithm.

## Data Availability

All data produced in the present study are available upon reasonable request to the authors

## AUTHOR CONTRIBUTION

BGS conceived and designed the study, conducted data analysis, interpreted the results, and drafted and revised the paper. PYY, CLC, MJS, RZ contributed to study design, data acquisition, results interpretation, and paper revision. RJS, CLC, RZ reviewed and further contributed to the manuscript. All authors drafted and revised the manuscript.

