## Supplemental Tables for "Development and Application of Pharmacological Statin-Associated Muscle Symptoms Phenotyping Algorithms Using Structured and Unstructured Electronic Health Records Data"

**Table S1:** SAMS-CI Score Breakdown and Clinical Scenarios

| **Score breakdown in each category** | **Clinical notes scenario** |
| --- | --- |
| A. location and pattern of muscle symptoms |  |
| Score: 3 | clinical notes explicitly mentioned bilateral muscle symptoms in larger muscles such as hips, legs, thighs, shoulders |
| 2 | bilateral symptoms in smaller muscles such as calves, arms |
| 2 | not explicitly mentioned bilateral symptoms but mentioned diagnostic terms such as myopathy, myositis and myalgia |
| 2 | bilateral mild symptoms such as muscle pain, muscle aches, cramps |
| 1 | only mentioned mild symptoms such as muscle pain, muscle aches, cramps but no mention of bilateral presentation |
| B. timing of muscle symptoms onset related to statin initiation |  |
| 3 | symptoms within 4 weeks of statin index date |
| 2 | symptoms between 4-12 weeks after statin index date |
| 1 | symptoms over 12 weeks of statin index date |
| C. timing of muscle improvement after statin withdrawal |  |
| 2 | symptoms improvement within 2 weeks after statin withdrawal |
| 1 | symptoms improvement between 2-4 weeks after statin withdrawal |
| 1 | no mentions of statin withdrawal or symptoms improvement |
| 0 | no symptoms improvement after 4 weeks |
| D. timing of muscle symptoms recurrence after second statin regimen |  |
| 3 | similar symptoms occurred within 4 weeks |
| 1 | similar symptoms occurred between 4-12 weeks |
| 1 | no mentions of rechallenge but patient has documented muscle symptoms for more than 2 types of statins |
| 0 | similar symptoms did not occur or occurred more than 12 weeks after |

**Table S2:** Case Vignettes for Case and Control Ascertainment

| **Scoring** | **Case 1** | **Case 2** |
| --- | --- | --- |
|  | DL is a 62 year old male who has a history of severe mitral regurgitation, severe pulmonary hypertension, coronary disease, status post stenting in June 2021, chronic systolic heart failure/ischemic cardiomyopathy, hypertension, hyperlipidemia, morbid obesity. Patient was discharged with atorvastatin 80mg on 6/20/2021.  On 07/01/2021, DL presented to the clinic complaining “aches and cramps” on atorvastatin 80mg. The provider recommended “hold of'' atorvastatin for 2 weeks.  On 7/28/2021, DL presents to the clinic again and states aching/cramping improved after atorvastatin hold. The provider initiated rosuvastatin 5mg for DL.  On 8/17/2021, DL presented to the clinic. DL reports that muscle symptoms returned and his rosuvastatin was stopped. | AB is a 54-year-old female who presents to the clinic today for a follow up visit regarding her dyslipidemia and some shortness of breath. Patient started simvastatin 20mg on 10/22/2012.  On 11/4/2012, AB presented to the clinic stating that she did develop myalgias and muscle weakness.  On 12/31/2012, AB presented to the clinic again and provider states the simvastatin was held for three to four weeks for which AB believes that her myalgias did improve and she is back to her baseline. |
| Location of symptoms score | 1 since pain not specific to any area | 1 since pain not specific to any area |
| Time of onset score | 3 since DL reports muscle aches within 4 weeks of statin initiation | 3 since AB reports muscle aches within 4 weeks of statin initiation |
| Time of improvement score | 1 since DL’s muscle symptoms improved within 2-4 weeks of atorvastatin | 1 because according to the provider, her muscle symptoms improved after 3 to 4 weeks of hold |
| Time of recurrence score | 3 since DL’s symptoms returned after initiation of rosuvastatin within 4 weeks | 1 because this information is unknown |
| Overall score and decision | 8, so DL was classified as SAMS case | 6, so AB was classified as not having SAMS |

**Table S3:** SAMS-related ICD9 and ICD10 Codes and Description

| **ICD Type** | **Code** | **Description** |
| --- | --- | --- |
| ICD9CM | 359.4 | Toxic myopathy |
| ICD9CM | 359.9 | Myopathy, unspecified |
| ICD9CM | 728 | Infective myositis |
| ICD9CM | 728.87 | Muscle weakness (generalized) |
| ICD9CM | 728.88 | Rhabdomyolysis |
| ICD9CM | 728.9 | Unspecified disorder of muscle, ligament, and fascia |
| ICD9CM | 729.1 | Myalgia and myositis, unspecified |
| ICD9CM | 729.5 | Pain in limb |
| ICD9CM | 729.82 | Cramp of limb |
| ICD10CM | G71.9 | Primary disorder of muscle, unspecified |
| ICD10CM | G72.0 | Drug-induced myopathy |
| ICD10CM | G72.2 | Myopathy due to other toxic agents |
| ICD10CM | G72.49 | Inflammatory myopathy NOS |
| ICD10CM | G72.81 | Acute necrotizing myopathy |
| ICD10CM | G72.9 | Myopathy, unspecified |
| ICD10CM | G73.7 | Myopathy in diseases classified elsewhere |
| ICD10CM | M60 | Myositis |
| ICD10CM | M60.0 | Infective myositis |
| ICD10CM | M60.00 | Infective myositis, unspecified site |
| ICD10CM | M60.003 | Infective myositis, unspecified right leg |
| ICD10CM | M60.003 | Infective myositis, right lower limb NOS |
| ICD10CM | M60.004 | Infective myositis, unspecified left leg |
| ICD10CM | M60.005 | Infective myositis, unspecified leg |
| ICD10CM | M60.005 | Infective myositis, lower limb NOS |
| ICD10CM | M60.009 | Infective myositis, unspecified site |
| ICD10CM | M60.01 | Infective myositis, shoulder |
| ICD10CM | M60.011 | Infective myositis, right shoulder |
| ICD10CM | M60.012 | Infective myositis, left shoulder |
| ICD10CM | M60.019 | Infective myositis, unspecified shoulder |
| ICD10CM | M60.05 | Infective myositis, thigh |
| ICD10CM | M60.051 | Infective myositis, right thigh |
| ICD10CM | M60.052 | Infective myositis, left thigh |
| ICD10CM | M60.059 | Infective myositis, unspecified thigh |
| ICD10CM | M60.06 | Infective myositis, lower leg |
| ICD10CM | M60.061 | Infective myositis, right lower leg |
| ICD10CM | M60.062 | Infective myositis, left lower leg |
| ICD10CM | M60.069 | Infective myositis, unspecified lower leg |
| ICD10CM | M60.162 | Interstitial myositis, left lower leg |
| ICD10CM | M60.8 | Other myositis |
| ICD10CM | M60.80 | Other myositis, unspecified site |
| ICD10CM | M60.81 | Other myositis shoulder |
| ICD10CM | M60.811 | Other myositis, right shoulder |
| ICD10CM | M60.812 | Other myositis, left shoulder |
| ICD10CM | M60.819 | Other myositis, unspecified shoulder |
| ICD10CM | M60.85 | Other myositis, thigh |
| ICD10CM | M60.851 | Other myositis, right thigh |
| ICD10CM | M60.852 | Other myositis, left thigh |
| ICD10CM | M60.859 | Other myositis, unspecified thigh |
| ICD10CM | M60.86 | Other myositis, lower leg |
| ICD10CM | M60.861 | Other myositis, right lower leg |
| ICD10CM | M60.862 | Other myositis, left lower leg |
| ICD10CM | M60.869 | Other myositis, unspecified lower leg |
| ICD10CM | M60.88 | Other myositis, other site |
| ICD10CM | M60.89 | Other myositis, multiple sites |
| ICD10CM | M60.9 | Myositis, unspecified |
| ICD10CM | M62.81 | Muscle weakness (generalized) |
| ICD10CM | M62.82 | Rhabdomyolysis |
| ICD10CM | M62.9 | Disorder of muscle, unspecified |
| ICD10CM | M79.1 | Myalgia |
| ICD10CM | M79.10 | Myalgia, unspecified site |
| ICD10CM | M79.18 | Myalgia, other site |
| ICD10CM | M79.60 | Pain in limb, unspecified |
| ICD10CM | M79.604 | Pain in right leg |
| ICD10CM | M79.605 | Pain in left lower limb NOS |
| ICD10CM | M79.606 | Pain in leg, unspecified |
| ICD10CM | M79.609 | Pain in limb NOS |
| ICD10CM | M79.65 | Pain in thigh |
| ICD10CM | M79.651 | Pain in right thigh |
| ICD10CM | M79.652 | Pain in left thigh |
| ICD10CM | M79.659 | Pain in unspecified thigh |
| ICD10CM | M79.66 | Pain in lower leg |
| ICD10CM | M79.661 | Pain in right lower leg |
| ICD10CM | M79.662 | Pain in left lower leg |
| ICD10CM | M79.669 | Pain in unspecified lower leg |
